# COVID-19 length of hospital stay: a systematic review and data synthesis

**DOI:** 10.1101/2020.04.30.20084780

**Authors:** Eleanor M Rees, Emily S Nightingale, Yalda Jafari, Naomi R Waterlow, Samuel Clifford, Carl A B Pearson, CMMID Working Group, Thibaut Jombart, Simon R Procter, Gwenan M Knight

## Abstract

**Background:** The COVID-19 pandemic has placed an unprecedented strain on health systems, with rapidly increasing demand for healthcare in hospitals and intensive care units (ICUs) worldwide. As the pandemic escalates, determining the resulting needs for healthcare resources (beds, staff, equipment) has become a key priority for many countries. Projecting future demand requires estimates of how long patients with COVID-19 need different levels of hospital care.

**Methods:** We performed a systematic review to gather data on length of stay (LoS) of patients with COVID-19 in hospital and in ICU. We subsequently developed a method to generate LoS distributions which combines summary statistics reported in multiple studies, accounting for differences in sample sizes. Applying this approach we provide distributions for general hospital and ICU LoS from studies in China and elsewhere, for use by the community.

**Results:** We identified 52 studies, the majority from China (46/52). Median hospital LoS ranged from 4 to 53 days within China, and 4 to 21 days outside of China, across 45 studies. ICU LoS was reported by eight studies - four each within and outside China - with median values ranging from 6 to 12 and 4 to 19 days, respectively. Our summary distributions have a median hospital LoS of 14 (IQR: 10-19) days for China, compared with 5 (IQR: 3-9) days outside of China. For ICU, the summary distributions are more similar (median (IQR) of 8 (5-13) days for China and 7 (4-11) days outside of China). There was a visible difference by discharge status, with patients who were discharged alive having longer LoS than those who died during their admission, but no trend associated with study date.

**Conclusion:** Patients with COVID-19 in China appeared to remain in hospital for longer than elsewhere. This may be explained by differences in criteria for admission and discharge between countries, and different timing within the pandemic. In the absence of local data, the combined summary LoS distributions provided here can be used to model bed demands for contingency planning and then updated, with the novel method presented here, as more studies with aggregated statistics emerge outside China.

## Background

As of the April 28th 2020, there have been over 3 million confirmed cases of COVID-19 and more than 200,000 deaths across 185 countries and territories [1]. Health systems are challenged by the influx of patients as SARS-CoV-2, the pathogen causing COVID-19, has spread throughout the world since its emergence in late December 2019 [2-6]. The risks of healthcare services being overwhelmed was most dramatically illustrated in Italy, where a rapid increase of COVID-19 cases needing hospitalisation pushed a well-equipped health system of 3.2 hospital beds per 1,000 people to breaking point [7]. This raises serious concerns over the potential impact on more resource-constrained health systems in low- and middle-income countries (LMICs) as epidemics begin to expand across Africa and South America.

Understanding and predicting hospital bed demand (as well as associated staff or equipment requirements) provides crucial evidence for decision-making and contingency planning (7,8). Predicting demand for hospital services requires an estimate of the number of patients requiring hospitalisation, and an estimate of how long each person will require hospital care. It is possible to model the rate of hospitalisation in many settings based on estimated epidemic curves. However, estimating length of stay (LoS) in hospitals requires observation of individual patient pathways.

COVID-19 presents at varying levels of severity. Hospital care can vary from general ward based care to high dependency units with oxygen support to intensive care where patients may be intubated for mechanical ventilation [8-10]. The LoS is likely to depend on the level of care required, as well as the geographic setting due to varying COVID-19 care guidelines. For example, some hospitals in China were initially used as isolation settings [11, 12]. As knowledge of effective treatments changes, the pathways, staff, beds and equipment required are also likely to affect the duration and level of care needed. Moreover, patient characteristics - such as age and comorbidities - impact disease severity [8,12-14] and are likely to influence LoS. If differences are significant then capacity planning may need to account for these characteristics to provide accurate predictions of the number of beds required at each level of care. Modelling studies predicting bed occupancy published so far have broadly relied on very few sources of information for LoS estimates, which were often derived from very different settings [15-22]. Estimates for LoS can be obtained from a variety of studies, but are often an incidental result rather than a study’s primary outcome, and typically only summary statistics are reported. In general, LoS distributions are right-skewed due to a minority of patients with long hospital stays and are often modelled using gamma, log-normal or weibull distributions [23] (although log-normal is less preferred due to its heavier tails). A particular challenge is how to synthesise appropriate LoS distributions from a range of relevant sources in similar settings, capturing the variation both within and between them. Incorporating the uncertainty and stochasticity in parameters using a distribution, rather than fixed point estimates (such as the mean over all studies), allows for more realistic model predictions.

We performed a systematic review to identify the current evidence on LoS for COVID-19 patients worldwide. We also present a method for generating LoS summary distributions by combining information from different summary statistics (mean and medians) reported in multiple studies, and accounting for differences in sample sizes. In doing this work, we aim to inform the efforts of modellers and policy makers to better anticipate healthcare needs during the evolving COVID-19 pandemic.

## Methods

### Search strategy

This study was conducted following the Preferred Reporting Items for Systematic Reviews (PRISMA) guidelines(9). We searched the bibliographic databases Embase and Medline, as well as the online preprint archive medRxiv. The latter was expected to be an important source due to the current rapid development of this field, hence the fully published literature would capture only a small proportion of the available information. We included articles published up to 2020-04-12 that reported a LoS for COVID-19 patients admitted to hospital. To ensure all relevant papers were captured, we examined the title, abstract and keywords of known studies reporting LoS to identify relevant search terms. Our search combined the concepts of COVID-19 (Coronavirus, COVID-19, 2019-nCov and SARS-CoV-2) with search terms related to duration of hospital stay (length of stay, admission duration, admission length, hospital*). The search terms for hospital stay length were kept broad to capture studies that report LoS as a secondary outcome. The full search terms for Embase, Medline and medRxiv are presented in the supplementary materials. In addition to our systematic searches, we also checked situation reports from the following organisations to see if they reported LoS estimates: UK Intensive Care National Audit and Research Centre (ICNARC); International Severe Acute Respiratory and Emerging Infection Consortium (ISARIC); World Health Organisation (WHO); the US Centers for Disease Control and Prevention (CDC); and China CDC and European CDC (ECDC).

### Inclusion and exclusion criteria

Inclusion criteria:

1. Studies that reported LoS in hospital for individuals who were admitted for confirmed COVID-19, or suspected COVID-19 which was later confirmed
2. Published (either as a pre-print or publication) between 2020-01-01 and 2020-04-12

Exclusion criteria:

1. Studies were excluded if LoS was reported for individuals only admitted to hospital for a reason other than confirmed or suspected COVID-19
2. Studies where the LoS endpoint was not death or discharge or continuing stay, for example transfer to another hospital
3. Studies which stated that hospitalisation was used as a form of isolation
4. Studies not published in English
5. Review articles

### Screening

The screening process is summarised in Fig. 1. All titles and abstracts were screened independently by two reviewers (EMR and SRP). Subsequently, abstracts and full texts of potentially relevant papers were independently reviewed by two reviewers (EMR and YJ).

**Figure 1.**
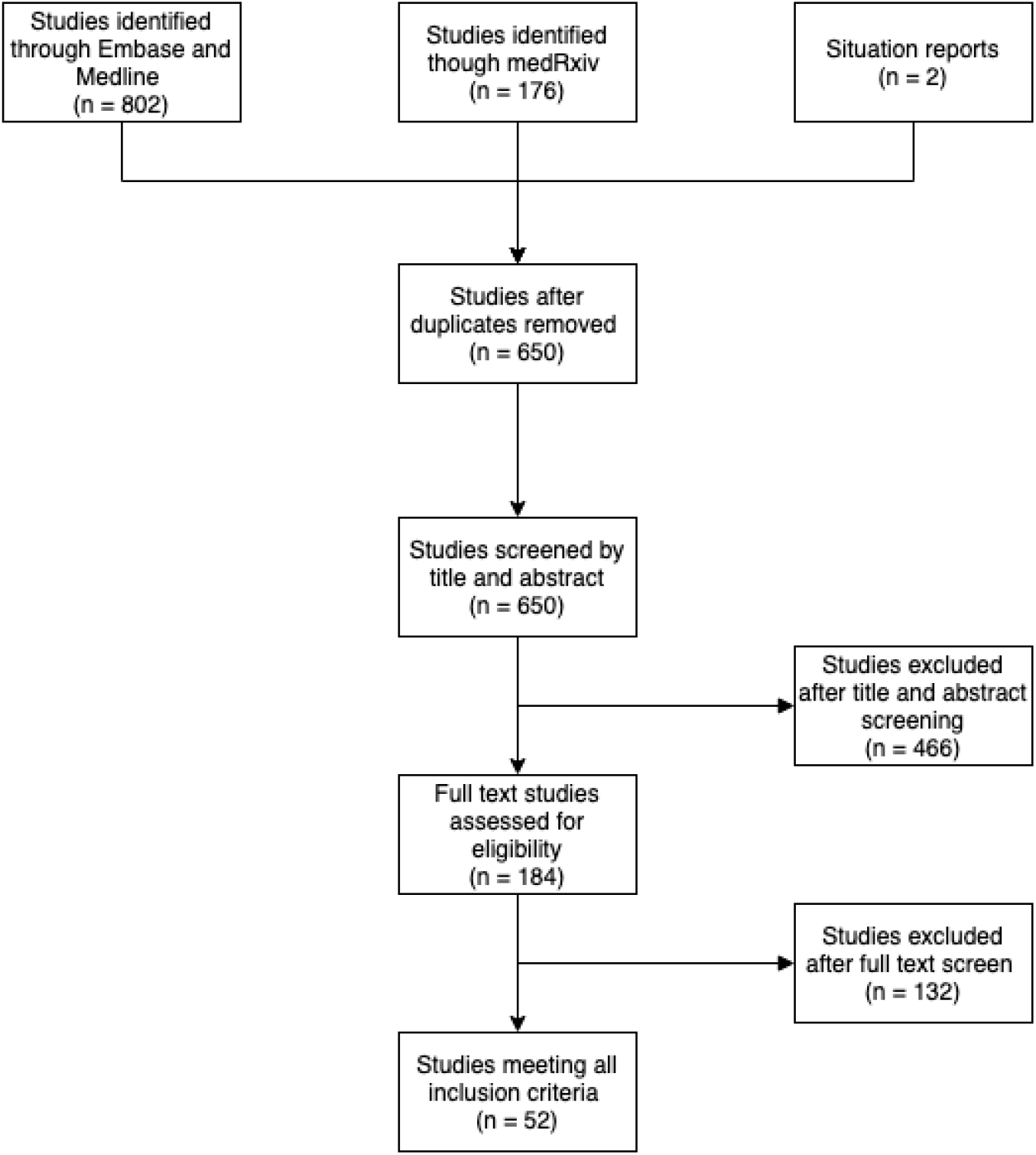
PRISMA diagram showing the results of the screening process used to identify included studies (n = 52).

### Data extraction and analysis

The data that was extracted from each study is presented in Supplementary Table A. Data extraction was performed by EMR, YJ and ESN, and then verified by a second member of the study team. Study characteristics (such as study dates, study population and study design) were recorded from each study, as well as information on the LoS sample estimate for both general hospital and intensive care units (ICUs), including sample size, discharge status and completeness of follow-up. If multiple LoS estimates were reported for different study populations these were all recorded (for example, LoS reported by disease severity, comorbidities and treatment groups). One study specified non-ICU LoS and this was grouped with general LoS estimates; ICU LoS was reported separately. Average patient age and sex distribution (% male) were summarised across all studies by weighted mean and standard deviation (mean (sd)), according to study sample size.

Where possible, LoS estimates were recorded as median and IQR. Otherwise, mean and SD, or in some cases only a point estimate was provided. Where estimates were presented as a mean, x, and standard deviation, s, we calculated the comparable quantiles from a fitted Weibull distribution by moment-matching using the *mixdist* package [24] and then discretising using the *distcrete* package [25] in R.

### Estimating LoS distributions

Overall summary distributions were created for general hospitalisation LoS and for ICU LoS. We included studies in the estimation of these summary distributions if they reported both the sample size along with either the median and interquartile range or the mean and standard deviation. If no measure of variation was provided (either IQR or standard deviation), the point estimates were included in figures but excluded from these summary results.

A Weibull distribution was fitted to the summary data from each grouping (by country setting and general/ICU classification) in the appropriate studies, using the Nelder-Mead method (using the *stats* package in R [26])for those reporting medians and IQRs, and the *mixdist* package [24] for those reporting means and standard deviations (as described in the previous section). The same approach was also tested using gamma distributions, but Weibull was marginally preferred with respect to total squared error in the fitted quantiles. These distributions were then discretised using the *distcrete* package in R [25]. 100,000 samples were then drawn from each of these distributions, with weighting according to their sample size. Specifically, the study distributions were first sampled according to a multinomial distribution defined by the studies’ relative sample sizes, and LoS was then sampled from each of these sampled distributions. Due to potential important differences in the characteristics of each study population, it may not be appropriate to weight entirely on sample size without considering how representative the cohort is of the general population. Therefore, as a sensitivity analysis, we performed the same analysis without weighting in order to understand how much this influences the distribution.

All analyses were performed using R version 3.6.3 (2020-02-29).

## Results

### Study characteristics

The results of our screening process are summarised in Fig. 1. After removing duplicates we found a total of 650 potentially eligible studies of which 52 studies met all the inclusion criteria. These included 32 peer-reviewed articles from the academic literature, 18 pre-print articles, and 2 reports from other sources ([27] and [28]). Several studies reported LoS by specific patient subgroups, according to disease severity, comorbidities (kidney injury, liver injury, hypertension, and cardiac injury), experimental treatments (heparin, lopinavir-ritonavir) and pregnancy status. A complete description of all reported LoS estimates are provided in Supplementary Table B. The key characteristics of the included studies are summarised in Table 1.

**Table 1:**
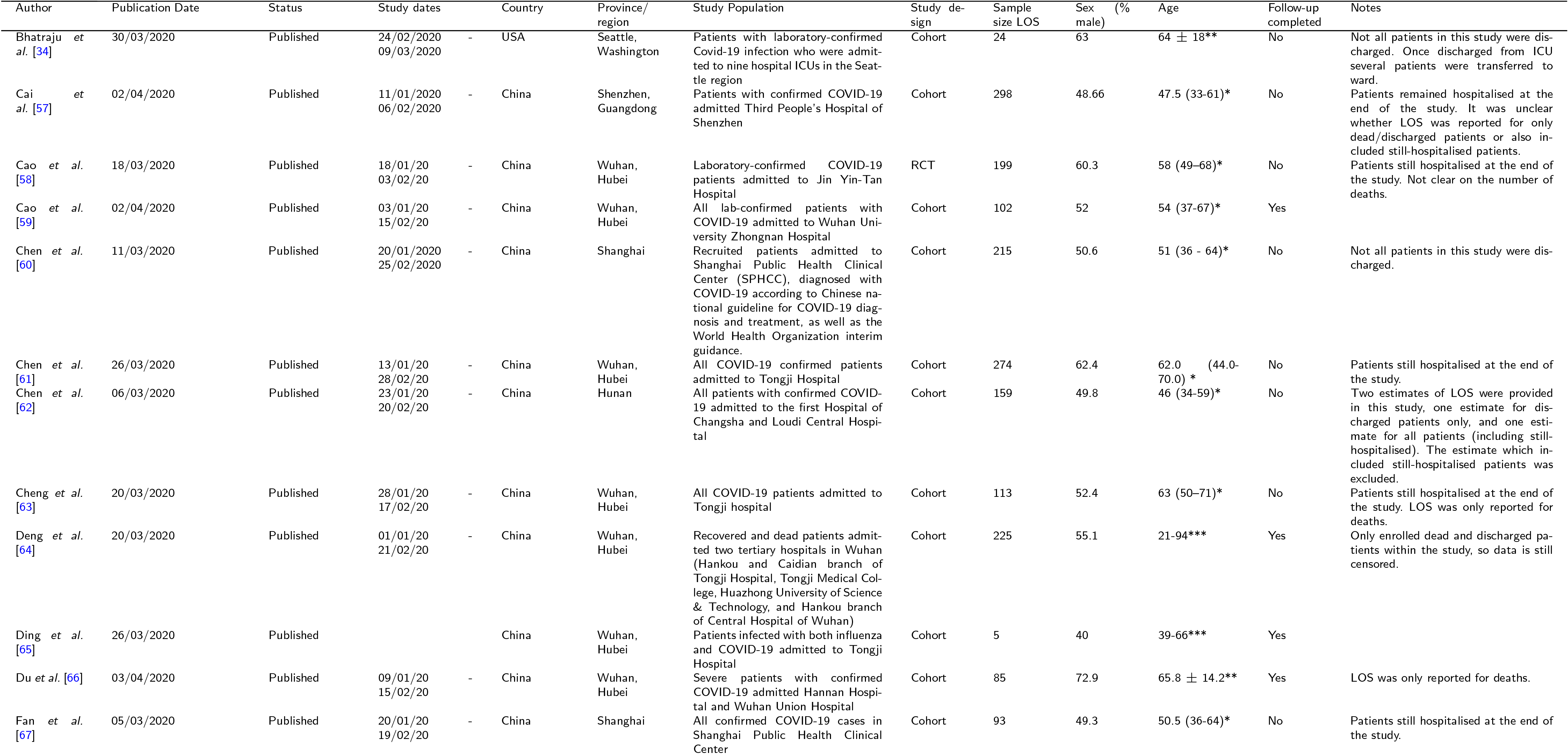
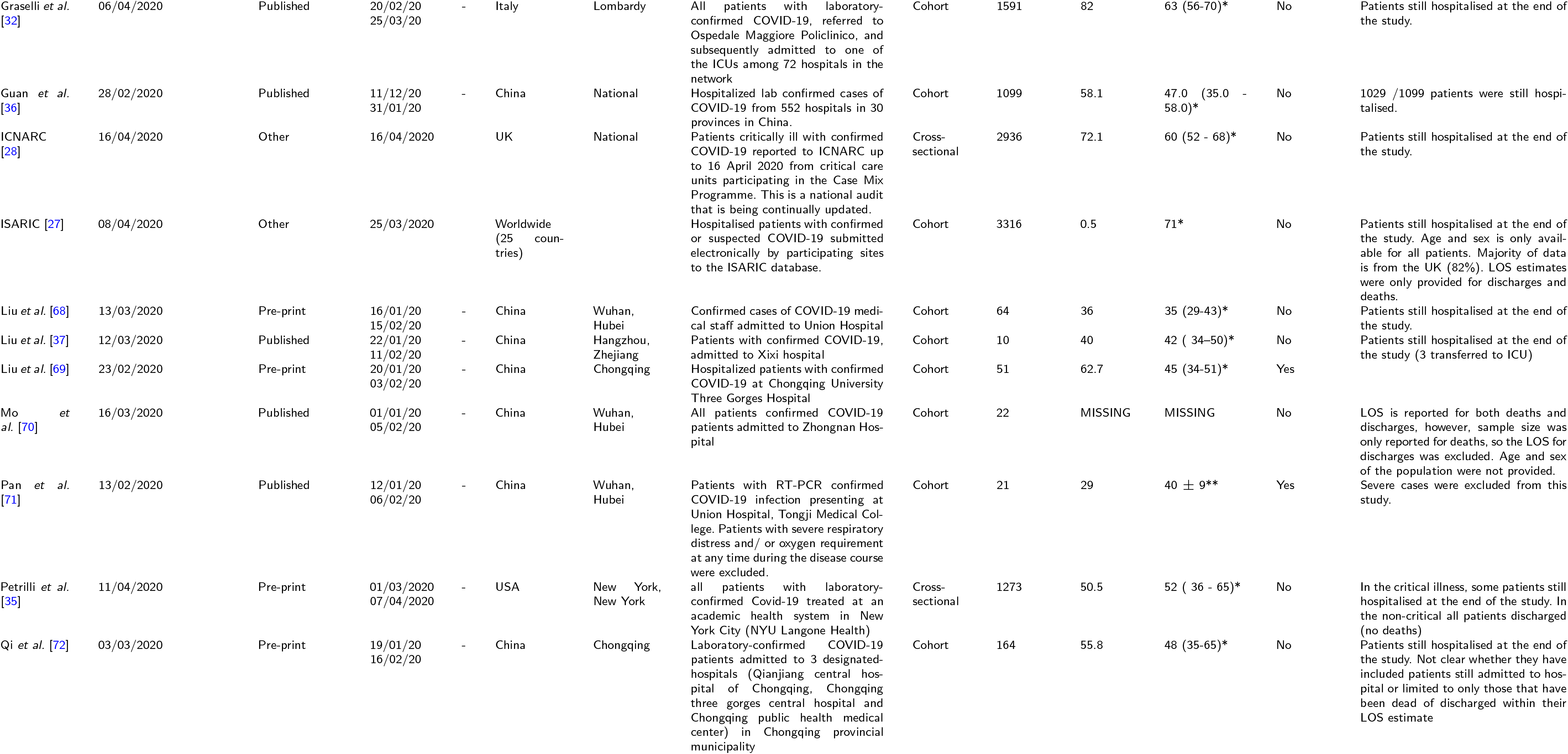
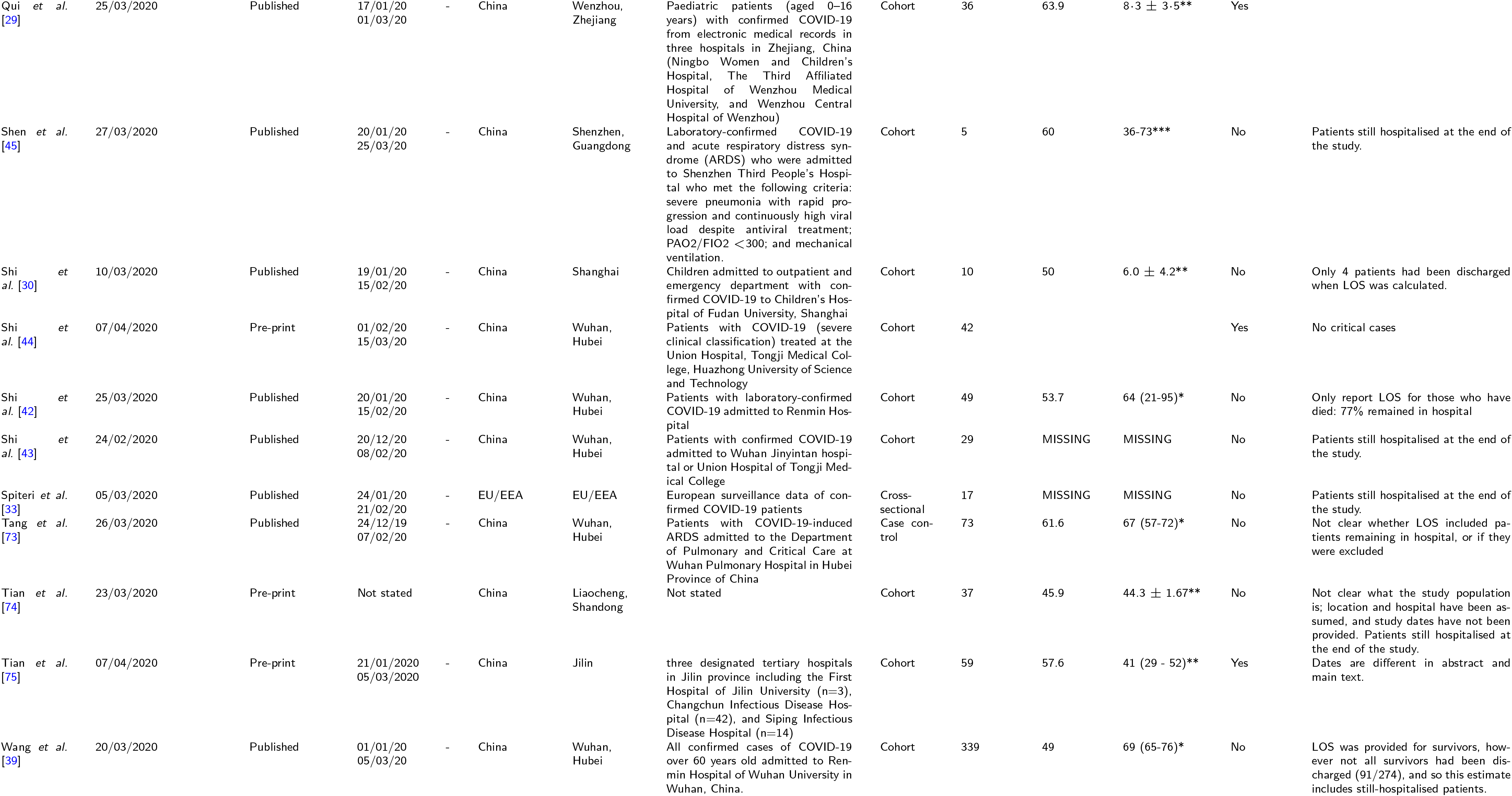
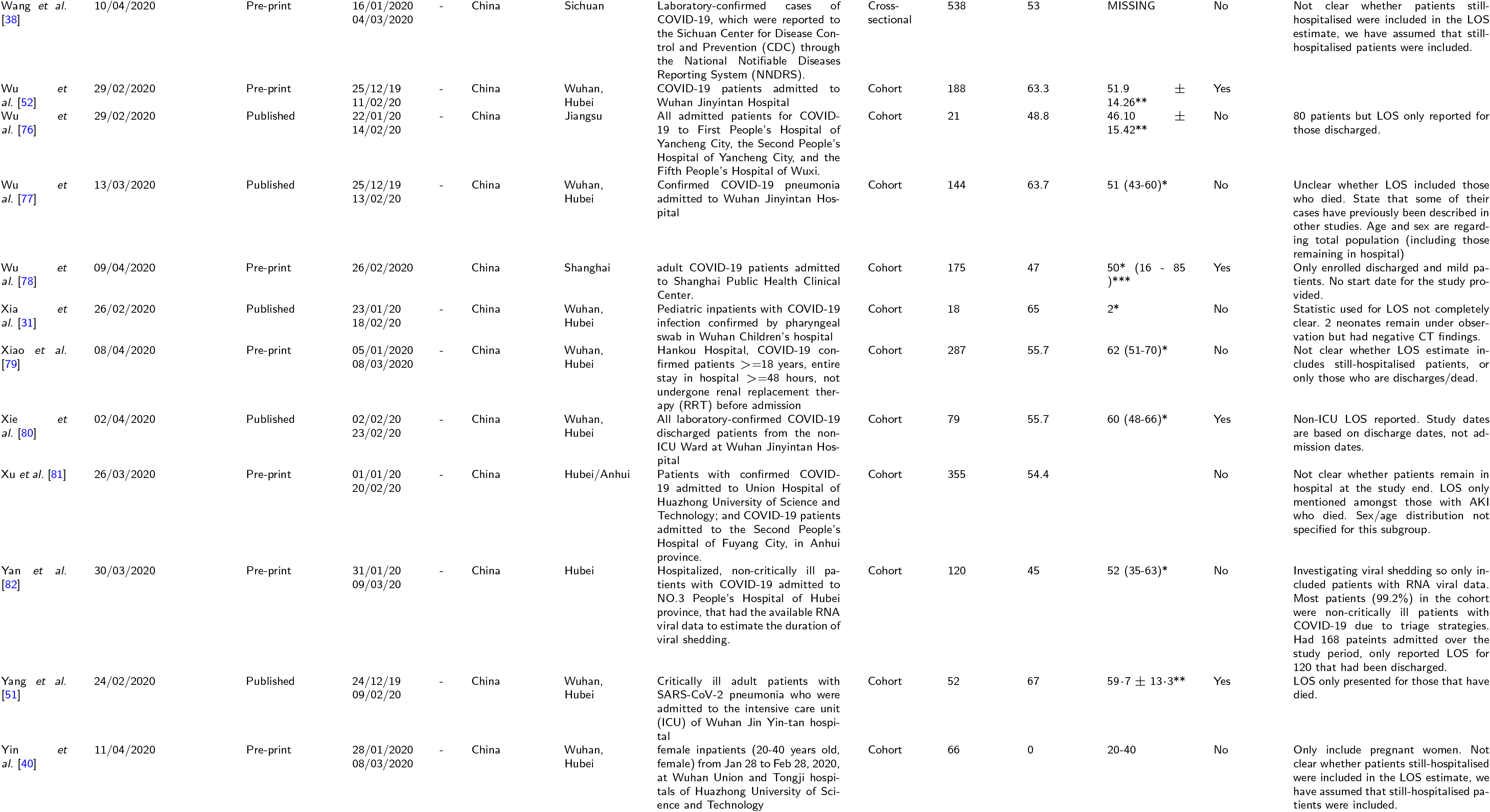
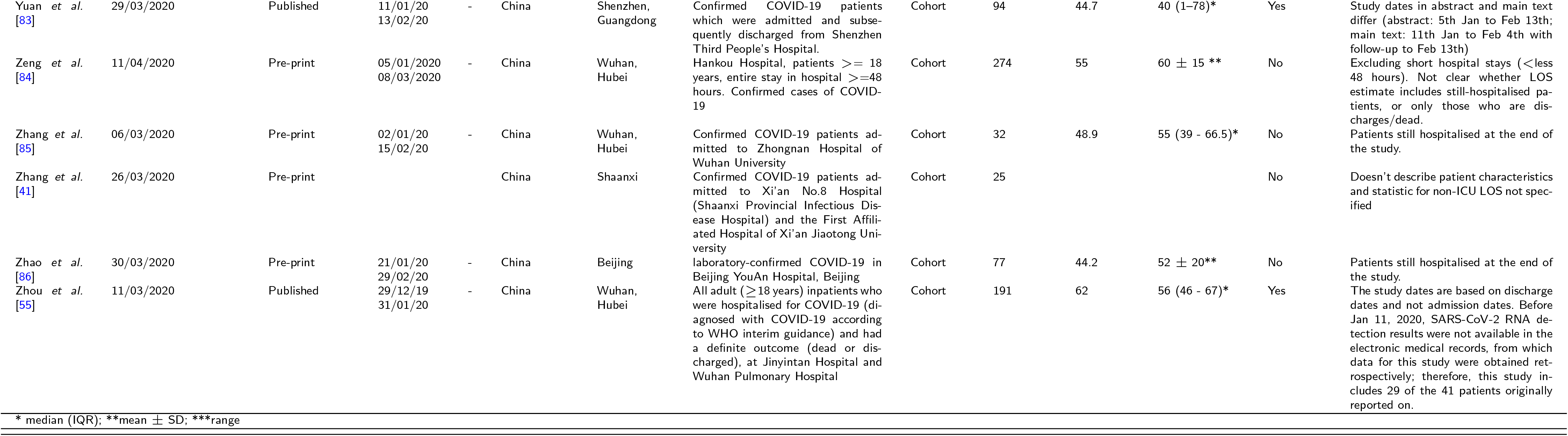
Summary study characteristics of included studies (n =52). A total of 46 studies were identified from China, and 6 studies identified outside of China.

The studies were carried out between 2019-12-24 and 2020-04-16. Although the cut-off was 2020-04-12 for inclusion of published and pre-print studies, the most recent version of the ICNARC report [28] was used, which included patients admitted up to 2020-04-16. The majority of studies were cohort studies (46/52), with four cross-sectional studies, one case-control and one randomised control trial (RCT). Two articles were reports from on-going data-collections (ISARIC [27], 2020-04-08, and ICNARC [28], 2020-04-16).

Studies were mostly conducted in adults with average participant age from 19 to 76 years (mean (sd) across studies, weighted by sample size: 59 (9.6) years), and overall reported only a slightly higher proportion of males to females (54 (10.9) % male). Three pediatric studies included patients from newborn to 18 years, with a weighted mean (sd) of 7 (2.8) years of age [29-31]. For the majority of studies LoS was a secondary or incidental outcome rather than the primary outcome. As a result, age and sex distributions were not always specific to the LoS population, and instead reported for the overall study population. Furthermore, it was not always possible to accurately interpret the sample size of the population, nor whether the LoS estimate included still-hospitalised patients. All LoS data extracted from studies are reported in Supplementary Table B.

The majority of the included studies (46/52) were based in China, with a particularly high number reported from Wuhan(27/46), and many study populations were from the same Outside of China, there was one study from Italy [32], one for the whole EU region [33], two from the United States [34,35], one from the United Kingdom [28] and one study that collated LoS estimates from multiple countries excluding China (although the majority of the data are from the UK; [27]).

Most studies (43/52) reported LoS for general hospitalisation only, with four studies reporting LoS for ICU only, and five studies reporting both. Only 15 studies reported LoS for study populations with completed follow-up (patient discharge or death), with 37 reporting estimates for populations where some patients remained in hospital or in ICU. The majority of studies only included discharged or dead patients within their LoS estimate, even if they had incomplete follow-up of the full cohort. However, for 8 studies it was unclear whether the reported LoS included patients who were still hospitalised [33,34,36-41].

### Overall hospital length of stay

Estimates of the overall hospital LoS are summarised in Fig. 2. Where provided, the overall study estimate of LoS for each discharge status is presented. For three studies, LoS was only reported within specific patient subgroups (relating to cardiac injury [42], COVID-19 recovery trajectory [43] and treatment comparison arms [44]), therefore in these cases we include both estimates. The longest stays were recorded in a study of five critically ill patients [45], of whom only three were discharged and all more than 50 days after admission, which does not appear representative of the overall distribution (see Fig. 2, Shen *et al*. (2020-01-20)). Excluding this study, the median duration of hospitalisation ranged from 5 to 29 days. There was no observed trend with respect to when the study was conducted (Fig 2).

**Figure 2.**
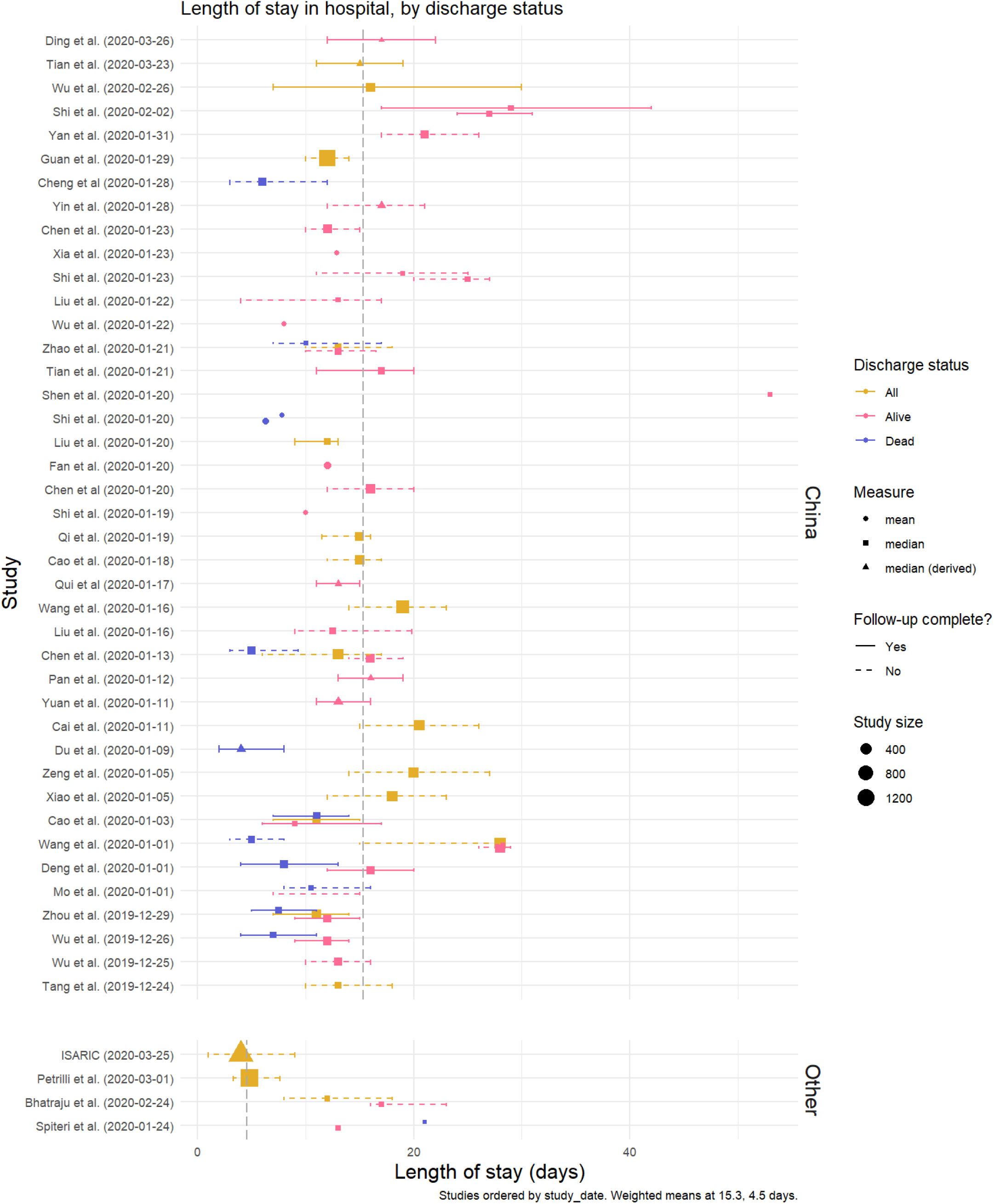
Hospital length of stay, by discharge status. Medians (square) are presented with interquartile range (IQR). Where estimates were reported as mean and standard deviation, equivalent quantiles have been calculated assuming a Weibull distribution (triangle); if no measure of variation was reported, only the original mean is presented (circle). The grey dashed lines represent the mean value across all point estimates within that setting, weighted by sample size.The studies are ordered by the study start date, with most recent at the top. Two studies (Shi et *al*. (2020-02-02) and Shi et *al*. (2020-01-23)) have multiple estimates for the same outcome which represent multiple treatment and comorbidity subgroups, respectively. Details of these are included in Table 1.

Estimates for LoS amongst patients who died in hospital were generally shorter than those for patients who were discharged alive, with medians between 4 and 21 days compared to 4 and 53 days, respectively. This difference is apparent in Fig. 2, where median LoS was lower for those discharged alive in 6 out of 8 studies that reported both outcomes. In studies that reported general hospital LoS by disease severity (11 studies, Supplementary Fig. A), there was a trend towards more severe cases having longer LoS. However, the definition of different levels of severity was inconsistent between studies so it is not possible to draw any confident conclusion.

Visual inspection of the study estimates suggested some evidence of a difference between general hospital LoS reported within and outside China, but studies outside China were too few (5/48) for a formal comparison. However, LoS reported within the ISARIC report [27] in particular (which includes contributed data from 25 countries, but with the majority of patients from the UK) gave a median and IQR (4 days (1 - 9)) substantially lower than the weighted mean from the studies from China (15.3 days).

The patient populations observed in these studies covered a wide range of ages, including three pediatric studies [29-31]. Among patients discharged alive there appears to be little difference in average LoS between studies with the youngest and oldest patients, but the longest estimates came from studies with average age in the upper end of the range (Wang *et al*. [39] and Shi *et al*. [44], with average age of 68 and 69, respectively; Supplementary Fig. B). The LoS estimates which included non-survivors tended to come from studies with older populations, as is to be expected given the well-documented, age-dependent fatality rate [46].

### ICU length of stay

Median stay in ICU ranged from 5 (IQR 2 - 9) to 19 (No IQR reported) days. There appeared to be less of a difference according to discharge status (alive or dead) than there was for general LoS (Fig. 3). A total of 8 studies reported ICU LoS estimates, with the same number of studies reporting LoS estimates from China and outside of China, and the resulting overall estimates are very similar. There were too few studies to conduct any comparison by age or disease severity.

**Figure 3.**
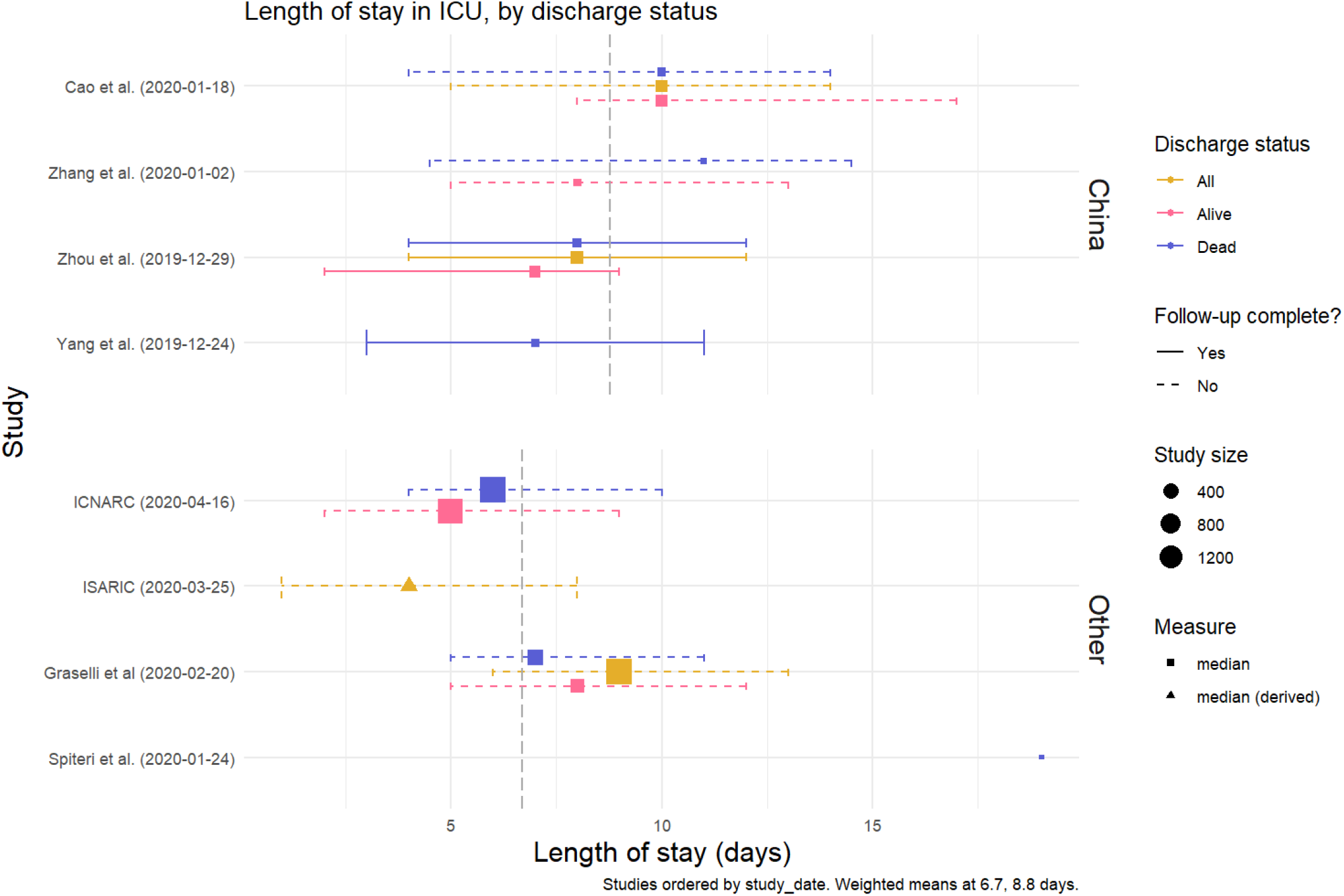
ICU length of stay, by discharge status. Medians (square) are presented with interquartile range (IQR). Where estimates were reported as mean and standard deviation, equivalent quantiles have been calculated assuming a Weibull distribution (triangle); if no measure of variation was reported, only the original mean is presented (circle). The grey dashed lines represent the mean value across all point estimates within that setting, weighted by sample size. Studies are ordered by the study start date.

### Estimated distributions

Estimated summary hospital LoS distributions for studies from China and studies outside China are shown in Fig. 4. The median and IQR for general hospital was estimated to be 14 (10-19) for China and 5 (3-9) excluding China. This was also repeated for ICU LoS, with a median and IQR 8 (5-13) for China and 7 (4-11) outside China. Studies from China which had complete follow-up with respect to general hospital LoS were compared with studies with incomplete follow-up. A slight difference was observed, with shorter median LoS observed in studies with complete follow-up (median, 12; IQR 8-17) compared with incomplete follow-up (median, 14; IQR, 10-19; Supplementary Fig. C). This was only performed for general hospital LoS in China, since no studies from outside China reported completed LoS for ICU.

**Figure 4.**
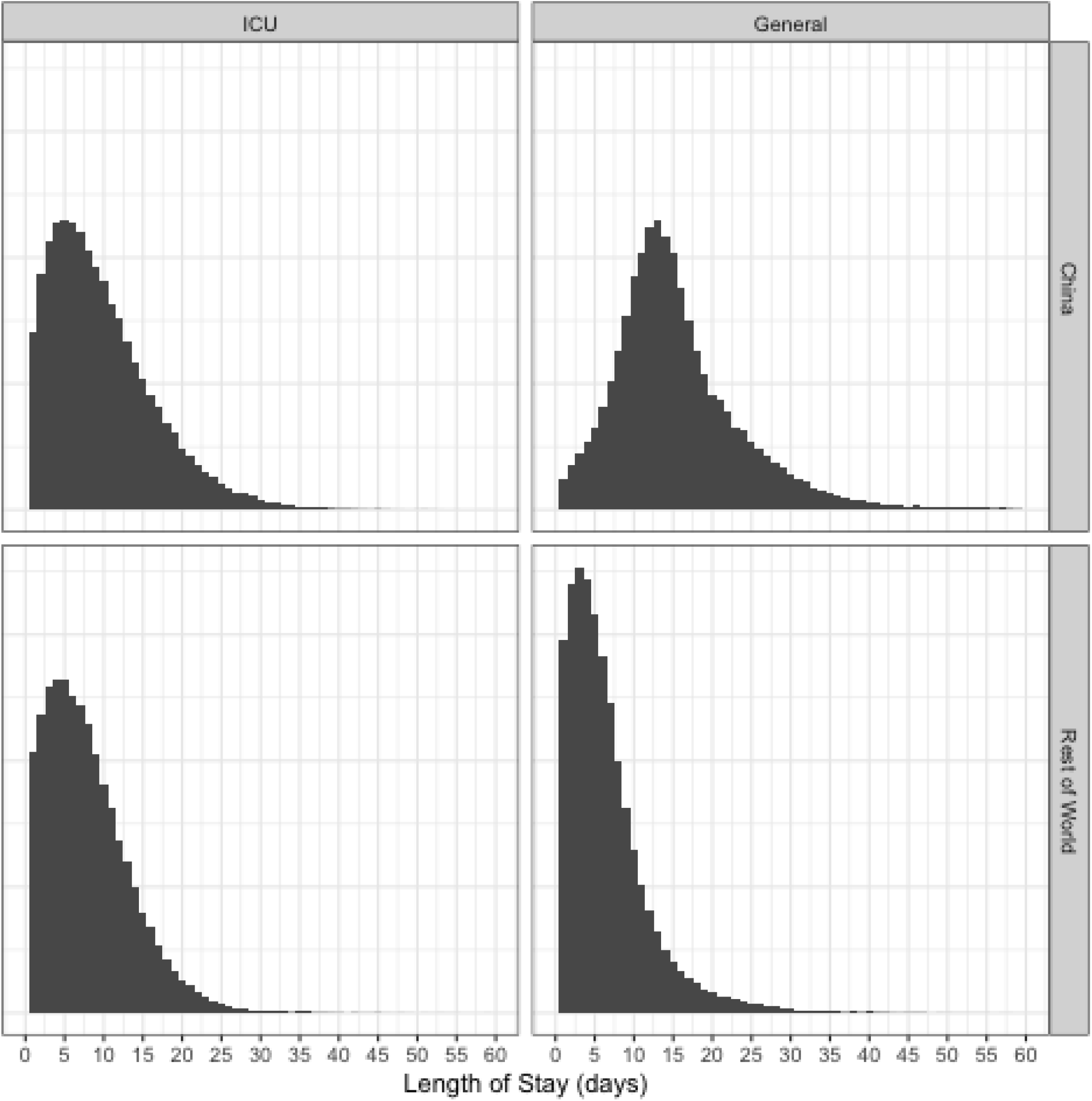
Combined LOS distributions. Samples from the overall LoS distributions, split by location (China or rest of world) and type (ICU vs General). For each subset, 100000 draws were taken. The x-axis was cut at days = 60.

For general hospital LoS in China, five studies were not included since they only provided point estimates for the LoS. The point estimates from four of these studies fell within the IQR of the estimated distribution, however, for [45], the point estimate was much longer. For general LoS outside of China, one study reported only a point estimate [33], and this also fell outside of the estimated IQR. Sensitivity analysis showed that weighting by sample size had minimal influence on the shape of these distributions (Supplementary Fig. D).

## Discussion

### Summary of findings

Understanding how long patients hospitalised with COVID-19 remain in hospital is critical for planning and predicting bed occupancy as well as associated staff and equipment needs. This review found that hospital LoS observations for COVID-19 patients published in the literature to date varied from less than a week to nearly two months. Stay in intensive care was shorter and less variable, with studies reporting medians of one to three weeks. Where LoS was reported according to discharge status, stay was found to be shorter for those who died than for those discharged alive; however, this difference was only apparent in terms of overall stay and not stay in ICU (no statistical comparison was made). With respect to practical implications, knowledge of a difference between survivors and non-survivors is of less use since the outcome will not be known in advance in order to influence decision-making. To the authors’ knowledge, this is the first formal review that has been conducted on hospital LoS for COVID-19.

The included studies yield some evidence of a difference between total hospital LoS observed in China and outside of China, with shorter LoS reported in the latter group (14 days (10-19) versus 5 days (3-9), respectively). However, only five studies were identified which reported LoS outside of China therefore this comparison is somewhat inconclusive. It may be that LoS is longer in China compared with other settings due to different criteria for hospital admission and discharge. A consensus exists across guidelines, such as ensuring resolution of symptoms and evidence of two negative PCR samples at least 24 hours apart before discharge [47,48], however, differences between settings may arise as a result of local capacity and strain on the health system. We attempted to capture this difference by recording time from onset of symptoms to admission, however, only one study outside of China reported this and a comparison was not possible. It is also possible that, with foresight from witnessing the Chinese epidemic, other countries set less strict criteria for discharge, in anticipation of stretched capacity. Other countries may also have used evidence from China to improve treatment methods and hence shorten LoS. However, this unfortunately appears unlikely as we did not observe a trend when looking at the reported LoS estimates over time.

In contrast, no difference was observed between settings for ICU LoS, for which there were an equal number of studies included from within and outside China. It is important to note that there might be key differences between ICUs in China compared with other countries, yet a definition for what constituted an ICU was rarely reported. Previous studies have found that ICU characteristics varied widely across geographic regions [49]. Further understanding of characteristics of ICUs reporting LoS for patients with COVID-19 are important in providing context on the reported estimates and should be investigated in future studies.

There appeared to be little difference of LoS observed by age in our results, apart from the fact that studies which reported deaths tended to have older patient populations. However, if there is indeed a trend we were unlikely to observe strong evidence for it amongst these studies, since the majority include a similar mix of ages, often tending towards older cohorts, and the age distribution was not always provided for the specific subgroup who had LoS recorded. Two studies [27, 38] were included in the review which reported LoS by age, and they both found longer LoS associated with older age groups. In addition, a study of LoS from the USA which was published after the search dates also reported a trend for longer LoS in older age groups [50].

### Limitations and biases

Having been the first country to observe this novel coronavirus, published data on COVID-19 patient outcomes in China is more widely available than from countries to which the epidemic spread later on. The set of studies found in this review reflects this bias towards evidence obtained from China, particularly Wuhan. As more studies emerge from a broader range of settings it would be important to re-evaluate LoS estimates, as there are likely to be between-country differences that we have not captured here.

Furthermore, a number of studies include patients from the same hospital over the same period, for example, Yang *et al*. [51] and Wu *et al*. [52] who both reported patients from Jin Yin Tan hospital in Wuhan), and it is possible that these studies had overlapping study populations. Furthermore, Guan *et al*. [36] was a national study conducted in China and ISARIC [27] included 25 countries world-wide, therefore these studies may also include patients previously described. The effect of this double-counting would be to bias the summary statistics towards the LoS from these settings. Although this is acknowledged as an issue, this was not considered as an exclusion criteria as it would have resulted in the exclusion of many studies. The overall benefit of inclusion was deemed to out-weigh the potential biases which may arise as a result of overlapping patient populations.

In this review we were only able to distinguish between “general hospital LoS” and “ICU LoS”, with many studies only reporting an overall LoS. This overall LoS will include both general hospital and ICU admissions within it. There is a need for more granularity with respect to patient pathways, distinguishing between admissions to different levels of care within one hospital episode in order to better inform healthcare contingencies. Patients may, for example, be transferred to ICU on more than one occasion during their stay, which is important to factor in when ICU capacity is particularly limited.

Changes in hospital demand may have also affected our estimates. At the beginning of the outbreak and in certain settings, hospitals were being used to isolate patients who were unable to isolate effectively at home [11,12]. This means that LoS for patients in some of the earlier studies within this dataset could have been longer due this logistical reason, rather than clinical need. Studies which mentioned this explicitly were excluded, yet there may still be others which were not so transparent. In addition, it is possible that, as hospitals reach the limits of their capacity, a more stringent triage policy may be implemented and the most critical patients may not be transferred to ICU. Despite this, we did not observe a trend when looking at the reported LoS estimates over time, suggesting that this is not in fact an important issue in our data.

Finally, many studies had incomplete follow-up with respect to LoS and, as a result, patients still hospitalised at the end of the study were not included in the summary statistics (right-truncation). This will bias estimates towards shorter LoS, as patients with longer LoS will not be included. A study by Lapidus *et al*. [53] investigated the bias associated with estimating average ICU LoS for COVID-19 patients based on observed LoS of discharged patients before follow-up of the entire patient cohort was completed. As expected, the authors found that the average LoS estimated at three months of follow-up was much longer than that estimated at one month. This potentially affects our estimates, given that 37 (out of 52) studies had incomplete follow-up with regards to LoS; although on comparison the difference between the groups was slight, and estimates where follow-up was complete were overall shorter. Several studies included still-hospitalised patients in their LoS summary without accounting for censoring [33,34,36-41], which potentially alters interpretation of the values.

### Summarising length of stay

We found that LoS is often not the primary measure of interest in studies which report it, however it is an important parameter when it comes to forecasting bed occupancy during an outbreak. By conducting this review we have systematically gathered a range of published estimates, providing a source from which researchers and decision makers can obtain estimates specific to their population of interest (e.g. with respect to comorbidities) and allowing comparison of LoS between several different populations and settings.

There have been numerous previous studies which have aimed to forecast the number of hospital beds required for COVID-19 patients [16-22,54]. Many of these studies published so far have used point estimates, only originating from one study which often does not reflect the context of interest. In particular, many used estimates from Zhou *et al*. [55] which reported a shorter general hospital LoS (median 110, IQR 7-0-14-0; Fig. 2), and a comparable ICU LoS (median 8-0, IQR 4-0-12-0; Fig. 3), compared with other studies from China. However, both of these were still longer than LoS estimates reported by studies outside of China. This means that the bed-forecasting studies relying on LoS estimates from Zhou *et al*. may be overestimating the number of beds required. The LoS estimate is a critical parameter within a bed forecasting model, and as such any model is likely to be very sensitive to the value or distribution being assumed, with huge implications for policy and planning.

This review has highlighted several potential sources of variation in LoS, and identified common issues and biases which influence each individual estimate. This gives a motivation for considering a wider range of values than can be obtained in a single study, aiming instead to capture the overall distribution of LoS across a variety of possible patient trajectories. Here we have included estimates of the overall LoS distribution in two settings (China/Other) for which we obtained sufficient data.

It is preferable to use data from the setting for which you are trying to forecast bed occupancy (as was done by the IHME COVID-19 health service utilization forecasting team [15]), however data on completed patient stays will often not be available until well after the onset of the epidemic. Furthermore, LMICs may have reduced capacity for surveillance and monitoring in order to obtain these data. In such cases, where countries are in the early stages of an outbreak, it would be better to use a conservative (i.e broad) distribution of LoS from another setting. As the pandemic progresses and more countries observe patients completing their hospital episodes, it will be possible to add further setting-specific summaries and improve this distribution.

As far as the authors are aware, the approach demonstrated here to summarise median and IQRs across multiple studies has not been proposed before, although there are similarities with the approach taken by others in the CMMID Working Group to pool *R_0_* estimates [56]. We present an intuitive method which exploits two optimisation methods to fit parametric distributions based on reported summary statistics rather than individual data, then samples across them. In this way we capture the central tendency and overall variation between a set of quantiles from different study populations. This allows multiple sources of evidence to be consolidated into a single distribution which can be used in bed forecasting going forward. By providing both the code for this analysis and our summary distributions, better bed occupancy predictions can be made in the future.

## Conclusion

This review summarised the available literature to provide estimates of LoS for general admission and ICU which can be applied for planning and preparedness for SARS-CoV-2. We found substantial differences between China and other settings in terms of total hospital stay, but little evidence for an impact on LoS of time of study, age or disease severity. We present summary distributions which can be used within models making predictions about bed requirements, and suggest that this may be a more robust and realistic way to characterise LoS than relying on summary data from just one setting or hospital. The majority of the data presented in this review comes from China and, as more data become available, it will be important to update this with setting-specific LoS estimates. Understanding the duration of hospitalisation of COVID-19 patients is critical for providing insights as to when hospitals will reach capacity, as well predicting associated staff or equipment requirements.

## Data Availability

All data extracted in this review and code used to analyse them are available via a github repository (https://github.com/esnightingale/los_review).

## Data and code availability

The data and code used in this work can be accessed at https://github.com/esnightingale/los_review.

## Author’s contributions

EMR - Conceptualization, Methodology, Literature search, Literature screening, Data extraction, Data analysis, Visualisation, Interpretation, Writing—original draft ESN - Methodology, Literature search, Literature screening, Data extraction, Data analysis, Visualisation, Interpretation, Writing—original draft YJ - Literature screening, Data extraction, Interpretation, Writing—review and editing NRW - Methodology, Data analysis, Visualisation, Writing—review and editing SC - Methodology, Writing—review and editing CABP - Code review, Writing—review and editing CMMID Working Group - Each contributed in processing, cleaning and interpretation of data, interpreted findings, contributed to the manuscript, and approved the work for publication TJ - Methodology, Code review, Writing—review and editing GMK - Conceptualization, Methodology, Interpretation, Writing—review and editing SRP - Conceptualization, Methodology, Literature screening, Data extraction, Interpretation, Writing—review and editing

## Acknowledgements

The authors gratefully acknowledge funding of the SPEAK India Consortium by the Bill and Melinda Gates Foundation (OPP1183986) for publication of this work.

We would also like to acknowledge the other members of the London School of Hygiene and Tropical Medicine COVID-19 modelling group, within the Centre for Mathematical Modelling of Infectious Diseases (CMMID working group), who contributed to this work: Rosalind M Eggo (HDR UK: MR/S003975/1, UK MRC: MC_PC19065), Megan Auzenbergs (BMGF: OPP1191821), Billy J Quilty (NIHR: 16/137/109), Nicholas G. Davies (NIHR: Health Protection Research Unit for Modelling Methodology HPRU-2012-10096), Mark Jit (BMGF: INV-003174, NIHR: 16/137/109, European Commission: 101003688), Jon C Emery (ERC Starting Grant: 757699), Petra Klepac (Royal Society: RP/EA/180004, European Commission: 101003688), Sam Abbott (Wellcome Trust: 210758/Z/18/Z), Sophie R Meakin (Wellcome Trust: 210758/Z/18/Z), Anna M Foss, Fiona Yueqian Sun (NIHR: 16/137/109), Stéphane Hué, Kathleen O’Reilly (BMGF: OPP1191821), Amy Gimma (Global Challenges Research Fund: ES/P010873/1), Adam J Kucharski (Wellcome Trust: 206250/Z/17/Z), Kiesha Prem (BMGF: INV-003174, European Commission: 101003688), Christopher I Jarvis (Global Challenges Research Fund: ES/P010873/1), Damien C Tully, Sebastian Funk (Wellcome Trust: 210758/Z/18/Z), David Simons (BBSRC LIDP: BB/M009513/1), Arminder K Deol, Alicia Rosello (NIHR: PR-OD-1017-20002), W John Edmunds (European Commission: 101003688), Joel Hellewell (Wellcome Trust: 210758/Z/18/Z), Timothy W Russell (Wellcome Trust: 206250/Z/17/Z), Graham Medley (BMGF: NTD Modelling Consortium OPP1184344), C Julian Villabona-Arenas (ERC Starting Grant: 757688), Kevin van Zandvoort (Elrha R2HC/UK DFID/Wellcome Trust/NIHR, DFID/Wellcome Trust: Epidemic Preparedness Coronavirus research programme 221303/Z/20/Z), Katherine E. Atkins (ERC Starting Grant: 757688), Quentin J Leclerc (UK MRC: LID DTP MR/N013638/1), Nikos I Bosse (Wellcome Trust: 210758/Z/18/Z), Yang Liu (BMGF: INV-003174, NIHR: 16/137/109, European Commission: 101003688), Akira Endo (Nakajima Foundation, Alan Turing Institute), Hamish P Gibbs (UK DHSC/UK Aid/NIHR: ITCRZ 03010), Charlie Diamond (NIHR: 16/137/109), Rachel Lowe (Royal Society: Dorothy Hodgkin Fellowship), Stefan Flasche (Wellcome Trust: 208812/Z/17/Z), Rein M G J Houben (ERC Starting Grant: 757699), James D Munday (Wellcome Trust: 210758/Z/18/Z).

## Supplementary materials

List of search terms

Supplementary Table A

Description of information extracted from included studies

Supplementary Table B

Database of all studies included within the review. Available as an excel spreadsheet: SupplementaryTableB.xlsx

Supplementary figures

A: LoS by disease severity

B: LoS by study median age

C: Summary distributions for China/other and general/ICU

D: Sensitivity analysis of summary distributions to weighting

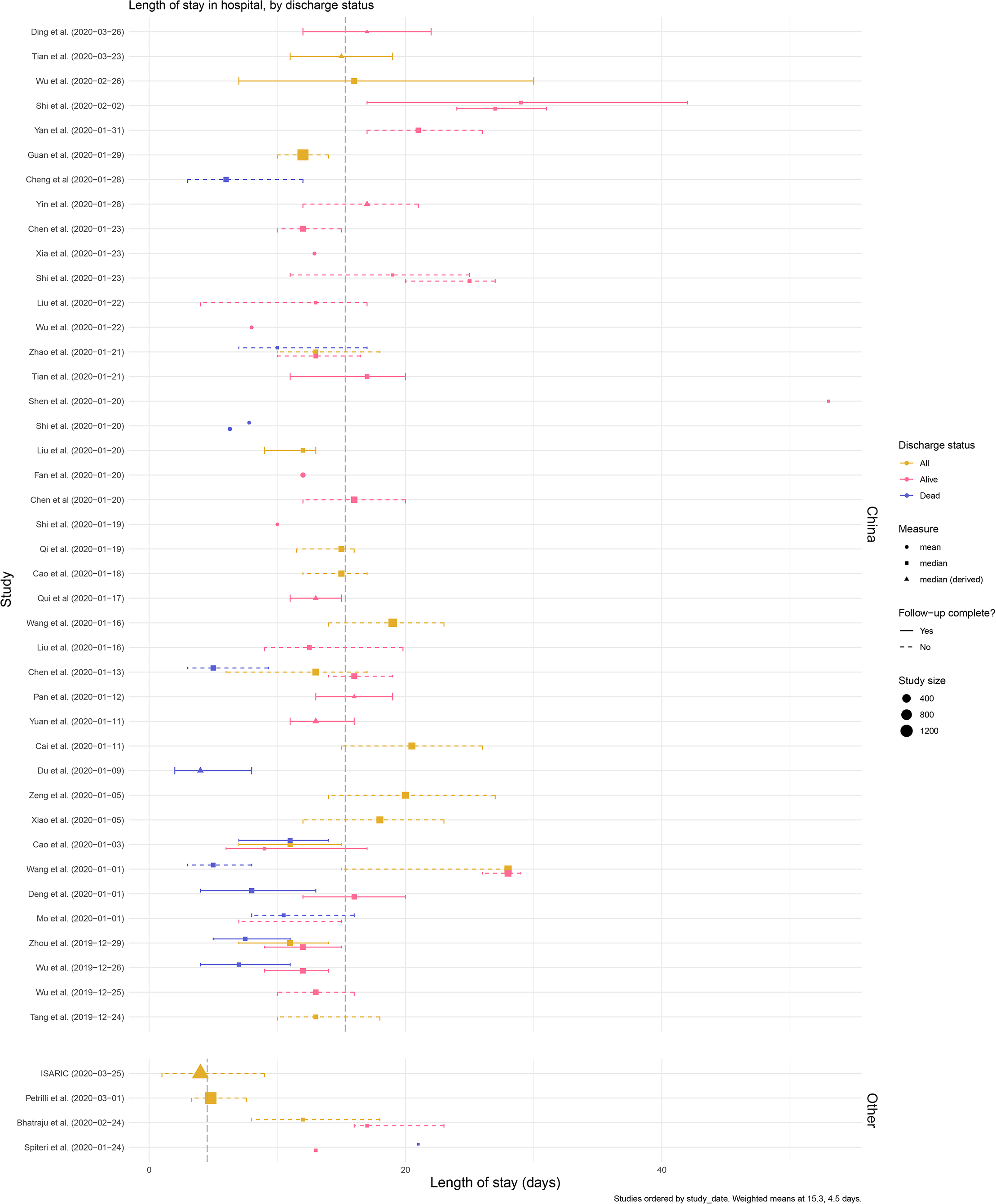

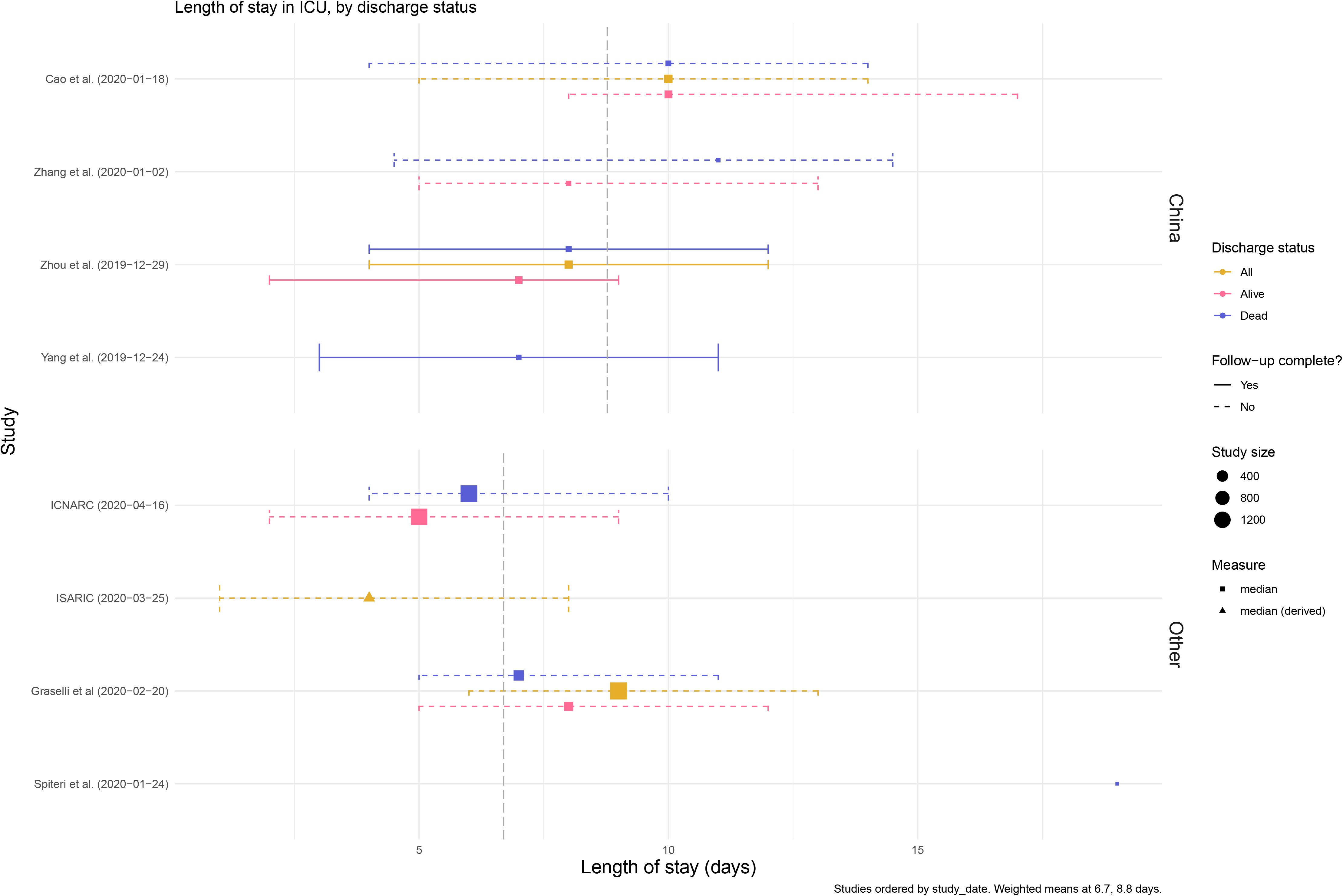

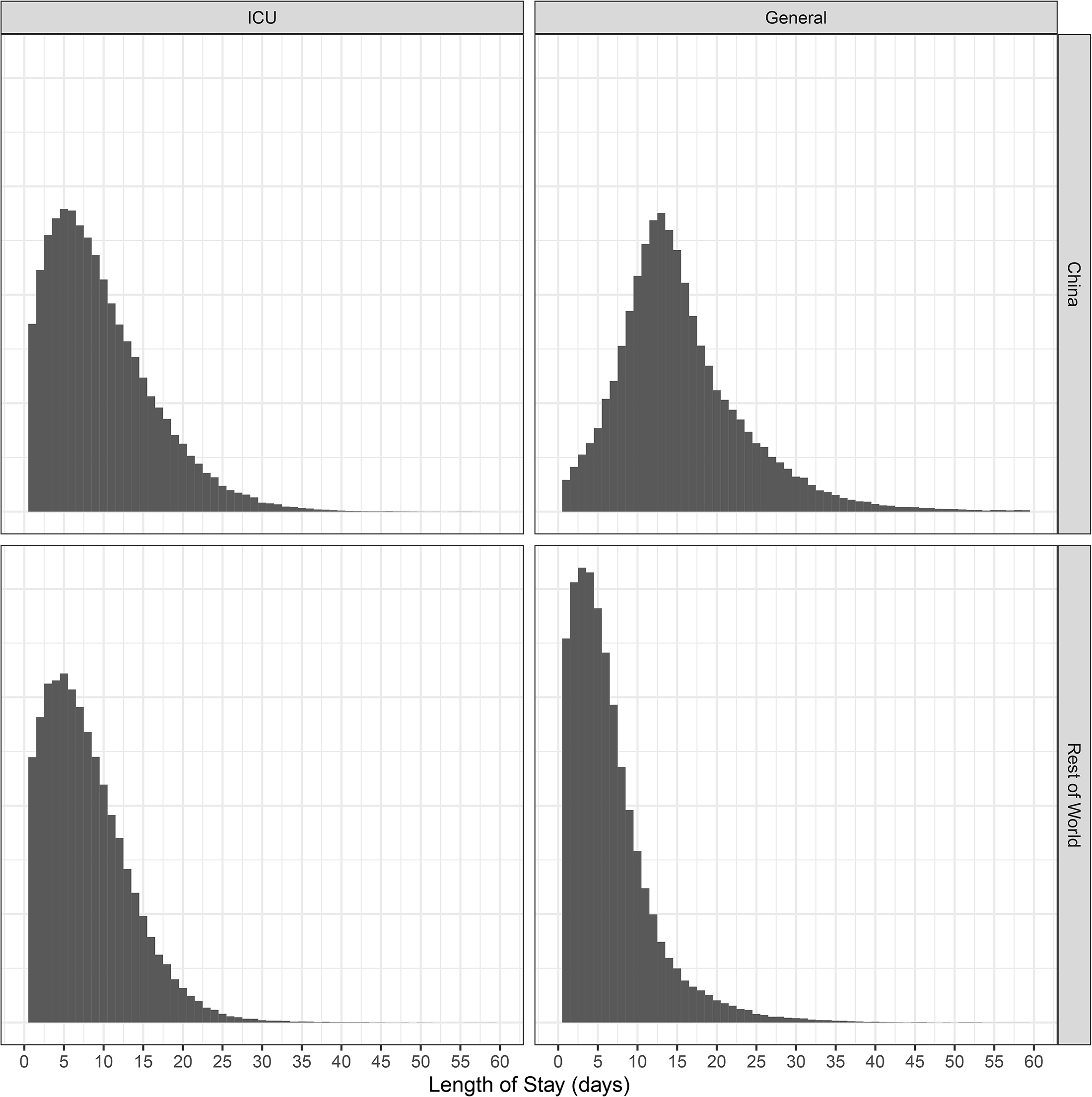

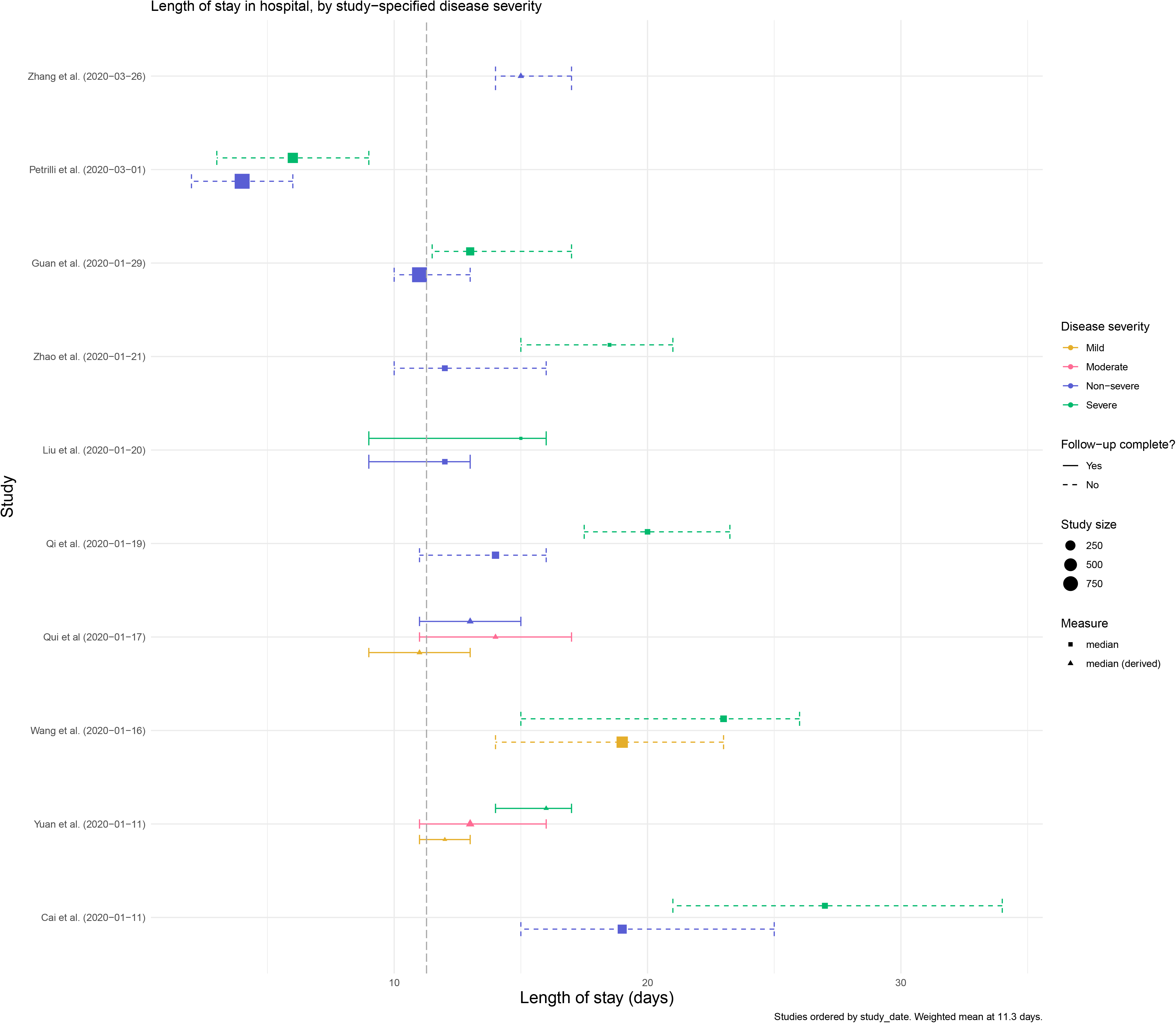

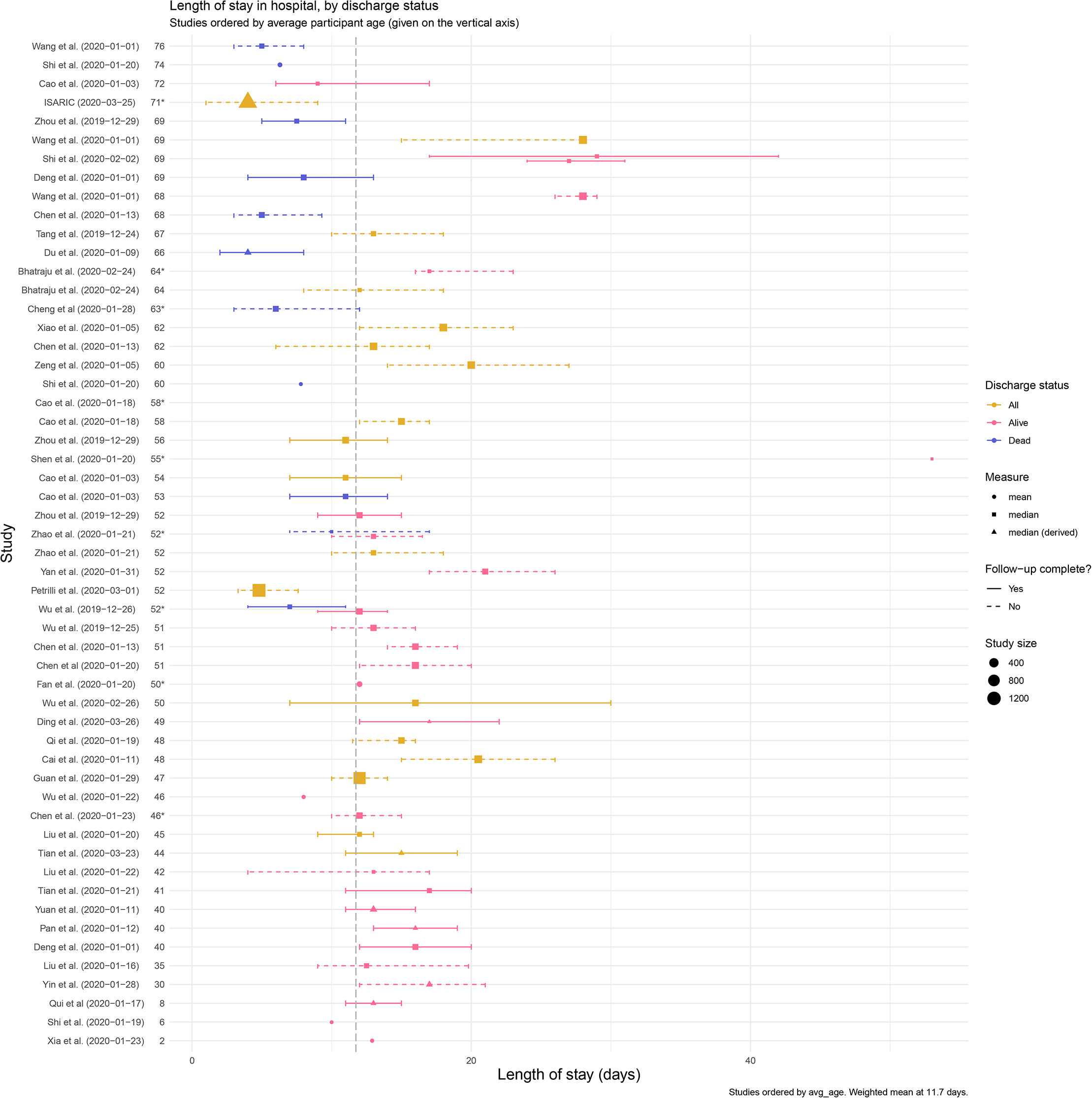

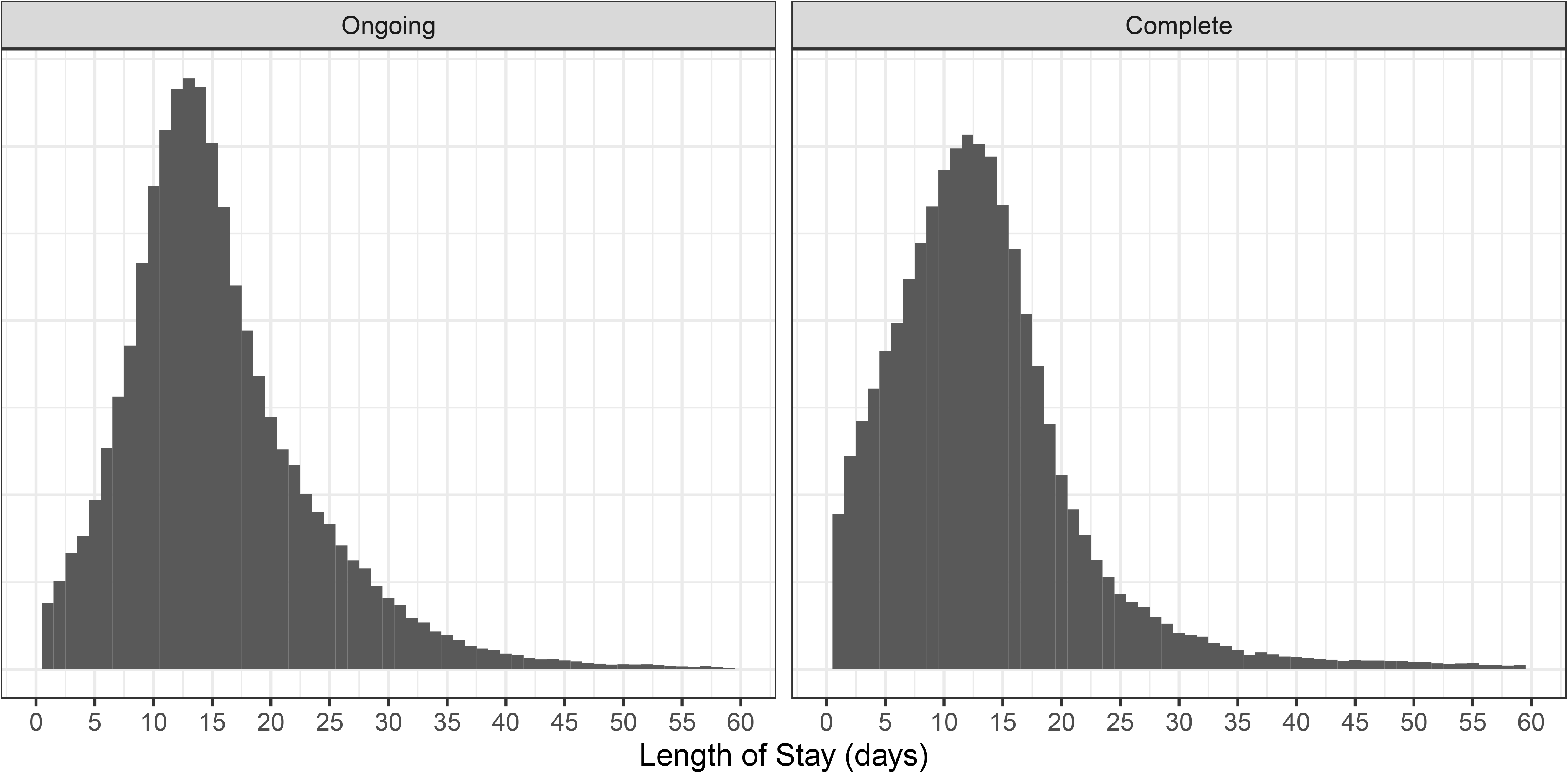

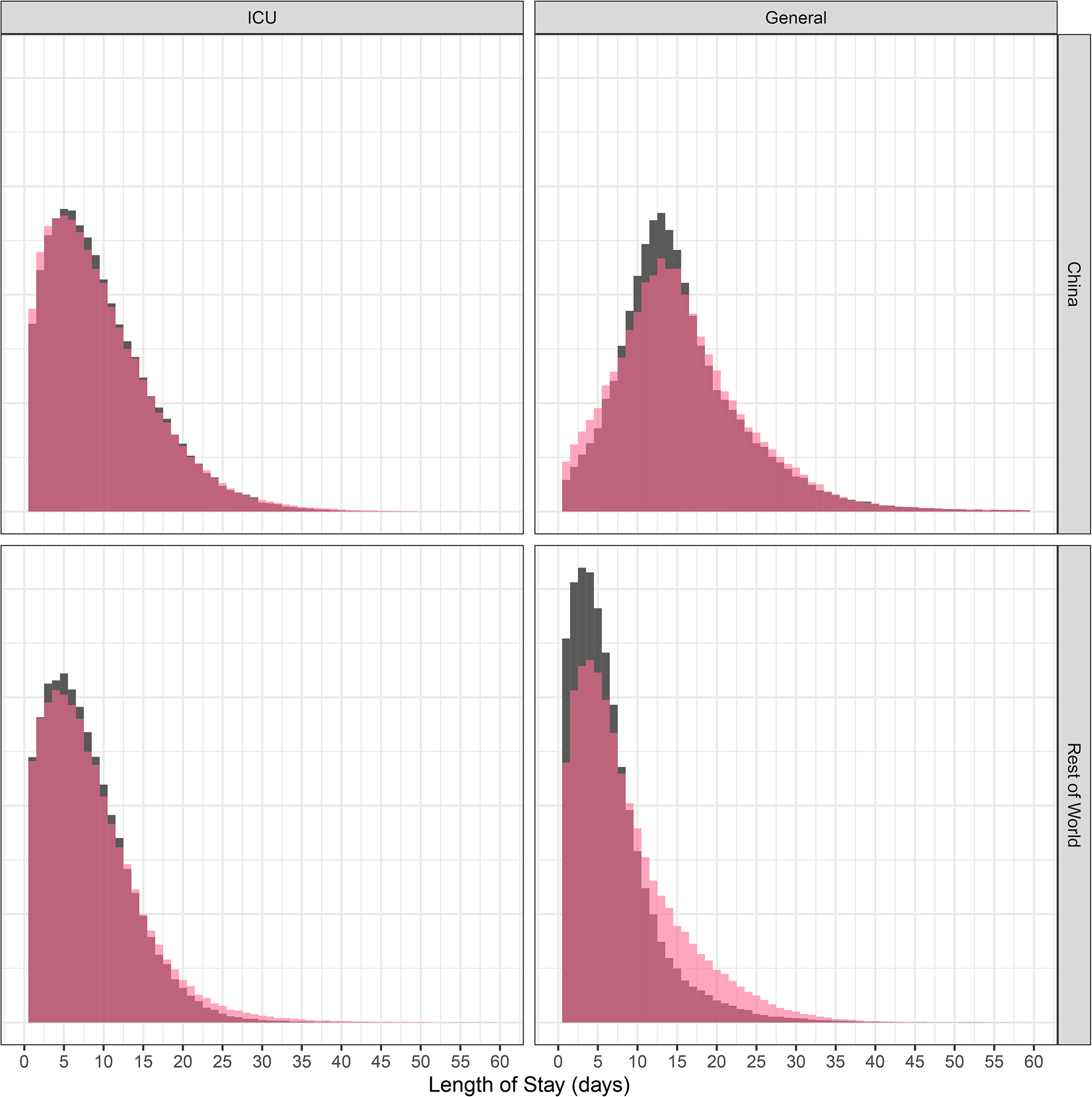

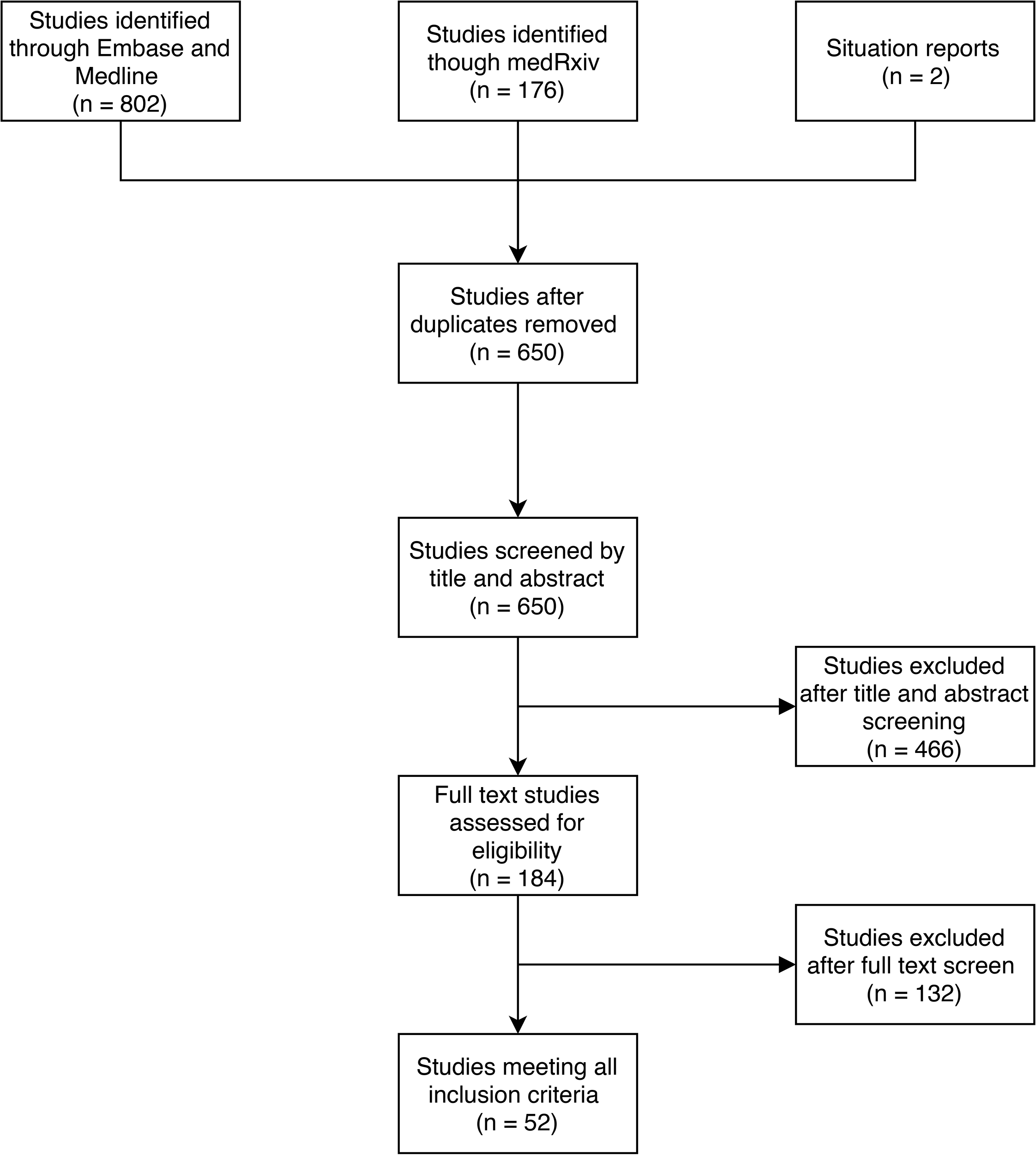

